# Mapping topography and network of brain injury in patients with disorders of consciousness

**DOI:** 10.1101/2022.08.22.22279090

**Authors:** Manoj Liyana Arachige, Udaya Seneviratne, Nevin John, Henry Ma, Thanh Phan

## Abstract

There is growing interest in the topography of brain regions associated with disorders of consciousness. This has caused increased research output, yielding many publications investigating the topic with varying methodologies. The objective of this study was to ascertain the topographical regions of the brain most frequently associated with disorders of consciousness.

We performed a cross-sectional text mining analysis of disorders of consciousness studies. A text mining algorithm built in the Python programming language searched documents for anatomical brain terminology. We reviewed PubMed studies up to 9^th^ July 2021 for the search query “Disorders of Consciousness.” The frequency of brain regions mentioned in these articles was recorded, ranked, then built into a graphical network. Subgroup analysis was performed by evaluating the impact on our results if analyses were based on abstracts, full-texts, or topic modelled groups (non-negative matric factorization was used to create subgroups of each collection based on their key topics). Brain terms were ranked by their frequency and concordance was measured between subgroups. Graphical analysis was performed to explore relationships between anatomical regions mentioned. The PageRank algorithm (used by Google to list search results in order of relevance) was used to determine global importance of the regions.

The PubMed search yielded 14,945 abstracts and 2178 full-texts. The topic-modelled subgroups contained 2440 abstracts and 367 full-texts. Text Mining across all document groups concordantly ranked the thalamus the highest (Savage score = 14.191). Graphical analysis had 4 clusters: cluster 1 had 20 members with the insular cortex [PageRank =0.167] as the most important member; cluster 2 had 29 members with the amygdala [PageRank =0.0199] being most important; cluster 3 had 10 members with the thalamus [PageRank = 0. 0205] being most important; cluster 4 had 19 members with the cingulate cortex [PageRank = 0. 0.020] being the most important.

The cingulate cortex and thalamus are strongly associated with disorders of consciousness, likely due to the roles they play in maintaining awareness and involvement in the Default Mode Network respectively. Other areas of the brain like the cuneus, amygdala and hippocampus should be further investigated.

## Introduction

Disorders of Consciousness (DoC) is a sequelae of traumatic brain injury (TBI) with an incidence of 0.7 per 100,000 TBI cases per year.[1, 2] Patients with DoC have impaired awareness and/or arousal and are often dependent on full-time nursing care in specialized wards and a multidisciplinary team of clinicians.[3] DoC have a profound impact on patients and their families.[4] The cost to the healthcare system, such as total inpatient cost of care, often exceeds $1,000,000 USD per patient [5, 6].

Diagnosis and prognosis of DoC remains challenging, in part from our incomplete understanding of this condition. There is a pervasive idea that DoC has a universally poor prognosis in the first 28 days; this has led to patients with DoC being labelled as a “neglected group of brain-injured patients”.[3, 7] A recent review on the subject drew attention to the different categories within the disorder and which subgroups may have late improvement.[7] Causative injuries can involve multiple areas of the brain and vary depending on the underlying etiology.[8-11] Individual studies, review or commentary articles on the topic have often discussed dysfunction in terms of large-scale functional networks such as the default mode network (DMN), and may refer to selected regions within the DMN[12-14]. Often these articles do not discuss the specific brain regions involved.[3, 15] As such, these papers do not provide practicing clinicians or those new to the topic with an understanding of the regions implicated in DoC and what they mean in terms of the brain imaging studies of patients under their care.[16, 17] We postulate that such data and its visual representation will be invaluable to clinicians to understand DoC in their patients.[18] This information may also assist future research such as targeted imaging studies in patients with DoC. In addition, the families of patients may want to know the nature of the imaging abnormalities and how this may fit into the concept of DoC. Such data may be used to better communicate this information to families.

Recognizing that only selected brain regions are mentioned in papers on DoC, we have opted not to perform a traditional meta-analysis but to take a crowd-sourcing approach. This takes the form of text mining literature on the topic to determine the brain structures involved in DoC. By text mining a large body of publications, the brain regions most commonly implicated in DoC according to the literature may be aggregated in an unbiased fashion.

## Methods

We performed a cross-sectional bibliometric (text mining) analysis of DoC publications. Because this article did not directly involve human subjects, while using only data from published articles, an institutional review board approval was not required. This study followed the Strengthening the Reporting of Observational Studies in Epidemiology (STROBE) reporting guideline.[19]

### Data Sources

We performed a search query for “Disorders of Consciousness” using the PubMed Entrez Programming Utilities.[20] Articles for the search query published up to 9^th^ July 2021 were retrieved. All studies from the initial search query were included. Two initial sets of texts (referred to henceforth as *corpora*) were created: the first corpus contained all abstracts from the PubMed e-search; the second contained all freely available full-texts from the PubMed e-search. Scripts for acquiring the abstracts and full texts can be found here: https://github.com/Mango117/BMedScDOC_Scripts

To allow for later subgroup analysis of the PubMed search query, a second, smaller pair of datasets were created from the two mentioned above. These subgroups were selected by using non-negative matrix factorization (NMF). NMF is an unsupervised method of clustering documents into “hidden topics.”[21] Using this method, relevant topics were filtered from the abstract and the full-texts corpora, then their documents separated for the subgroups (Fig 1).[22] Thus, 4 main datasets were created for analysis: an abstracts corpus; a full-texts corpus; a subgroup corpus which comprised of selected abstracts based on their NMF topics; and a final corpus of selected full-texts based on NMF topics. The code used to perform this NMF can be found here: https://github.com/Mango117/BMedScDOC_Scripts/tree/main/Topic_Model

**Fig. 1:**
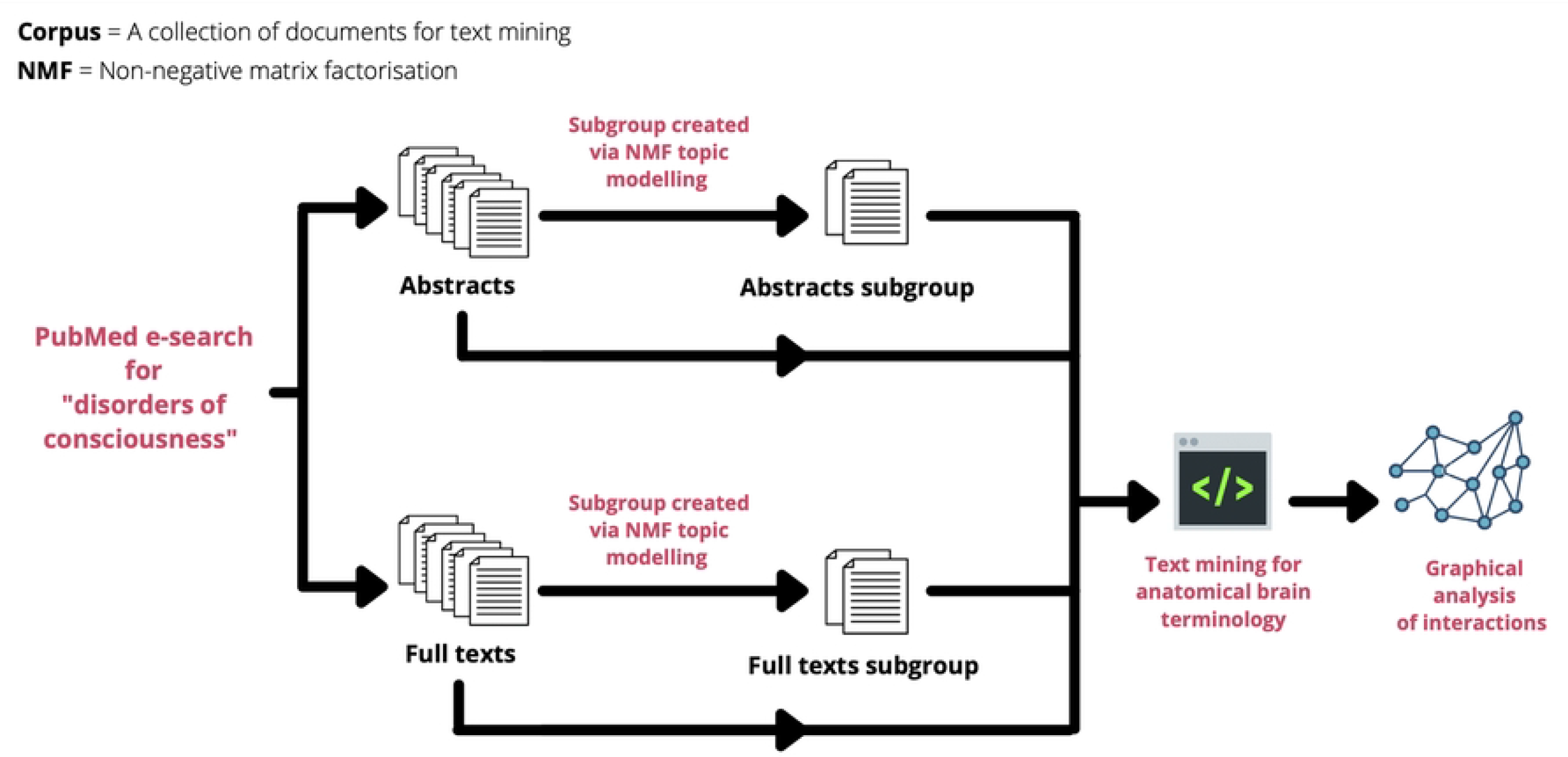
Flowchart of how each corpus was prepared from the original PubMed search query.

### Anatomical Dictionary

To perform the bibliometric analysis, an anatomical dictionary of brain terms fit for text mining was created using previously established neuroanatomical libraries. These terms would be used to search for keywords within the corpora of PubMed documents. We opted to combine two well-established catalogues of brain regions for the search. This would maximize coverage of cortical areas while allowing for varying neuroanatomical terminology. The terms were sourced from the automated anatomical labelling atlas 3 (AAL3) which lists 170 discrete regions of the brain, as well as the widely used Brodmann areas which separate the brain into 52 distinct histological areas.[23] This combined anatomical dictionary was modified slightly by removing overlapping search terms and making some linguistic changes to optimize the accuracy of the text mining process (Table 1).

**Table 1:**
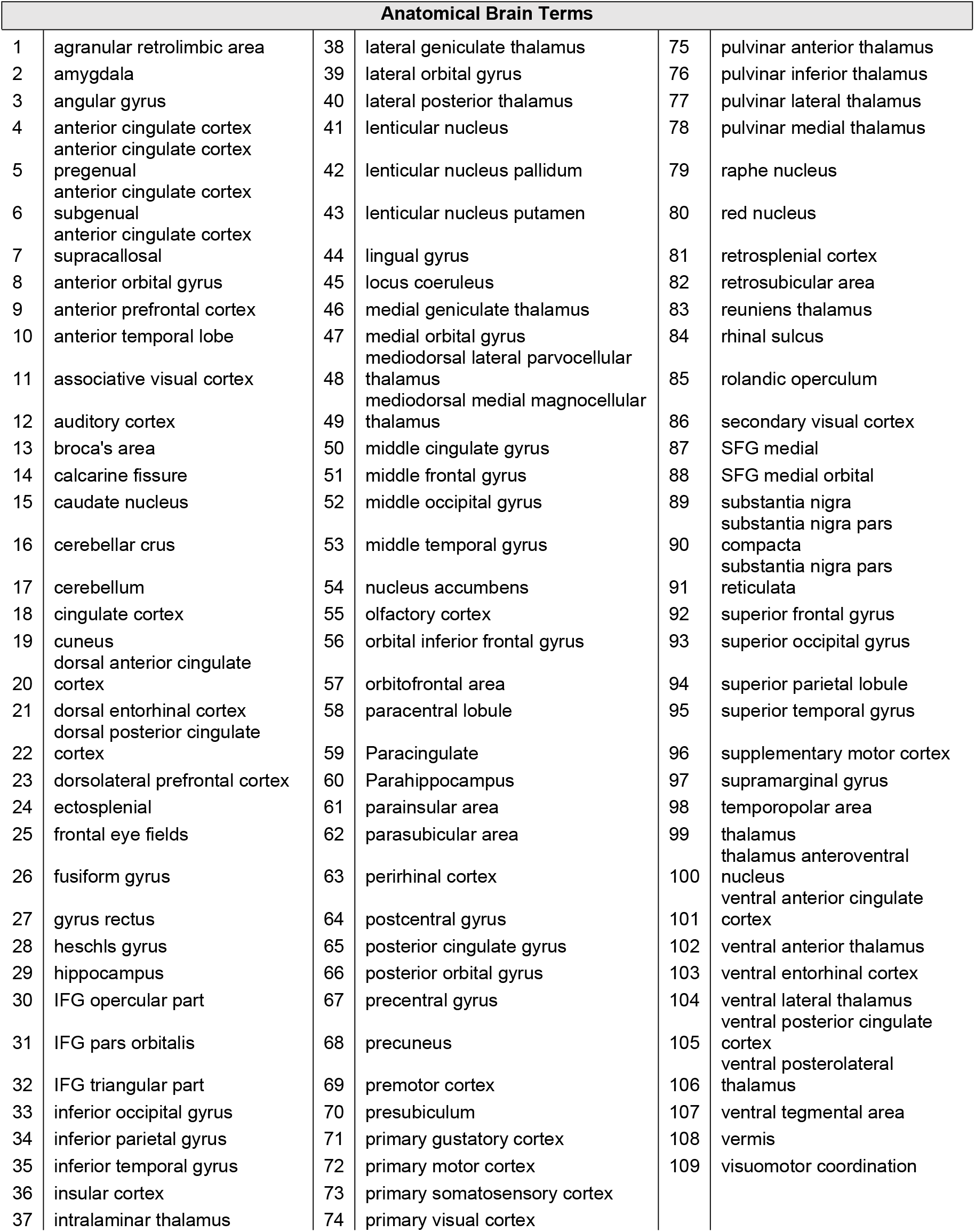
Combined and abbreviated brain dictionary of 109 anatomical brain regions created from AAL3 Anatomical Atlas and Brodmann Areas. Arranged in alphabetical order.

### Text mining

After establishing the corpora of documents and the dictionary of anatomical terms, text-mining analysis was performed on all four corpora. Each document was pre-processed to clean data for text mining.[24] After pre-processing, a term-document matrix (TDM) was created by calculating the frequency of each term from the anatomical dictionary in each corpus.

### Statistical Analysis

Statistical analysis was performed with Python (Python Software Foundation; version 3.9.5, https://www.python.org/).[25] The frequency of dictionary terms in each corpus was first ranked. To deduce a combined overall ranking across the entire data set, we calculated the concordance of rankings between the four corpora using Kendall’s rank correlation coefficient. Next, we used Savage Scores to assign an exponential weighting scheme to ranking. This ensures that the most important terms receive a higher weighting when concordance between datasets is calculated with Kendall’s rank correlation coefficient. This top-down approach to concordance is relevant in this study as brain areas ranked 1^st^, 2^nd^ and 3^rd^ should be weighted more than those placed 20^th^, 21^st^ and 22^nd^. The final Kendall’s value computed on Savage scores will be between 0 and 1, with 0 indicating no concordance and 1 indicating full concordance.[26, 27]

In addition, the full-texts corpus was selected for further graphical analysis using R (The R Foundation for Statistical Computing; version 4.1.0) to explore the associations between brain regions. The network manipulation software, Gephi (Gephi 0.9.2), was used for visualization of the data (including cluster analysis and calculation of a PageRank value to indicate the relative importance of each dictionary term in the graph)[28-30].

## Results

The PubMed search yielded a total of 14,945 results for the query “Disorders of Consciousness.” Data extraction garnered all 14,945 abstracts and 2178 full-text studies that were freely accessible for download. The abstracts and full-texts acquired were allocated into separate corpora. Analysis of the corpora revealed that the mean wordcount was 137.7 [95% CI, 136 to 139] for the abstract corpus and 2420.2 [95% CI, 2350 to 2490] for the full-texts corpus. NMF of the abstracts uncovered 30 topics, and 5 of these topics were selected as being most pertinent to DoC, yielding a subgroup of 2440 abstracts. NMF of the full-texts corpus uncovered a total of 20 topics, with 3 selected for the full-texts subgroup. This produced 367 documents.

Notably, the thalamus was the most frequent dictionary term across all corpora. It had 4050 mentions across the 4 datasets and was the highest ranked in each one (Table 2). The 4 corpora were considered significantly coherent with a Kendall’s Tau computed on Savage scores of 0.8380 (p < 0.001). Globally, the thalamus was ranked the highest (Savage score = 14.191), followed by the cingulate cortex (Savage score = 8.024) and the hippocampus (Savage score = 6.523) (Table 3).

**Table 2:**
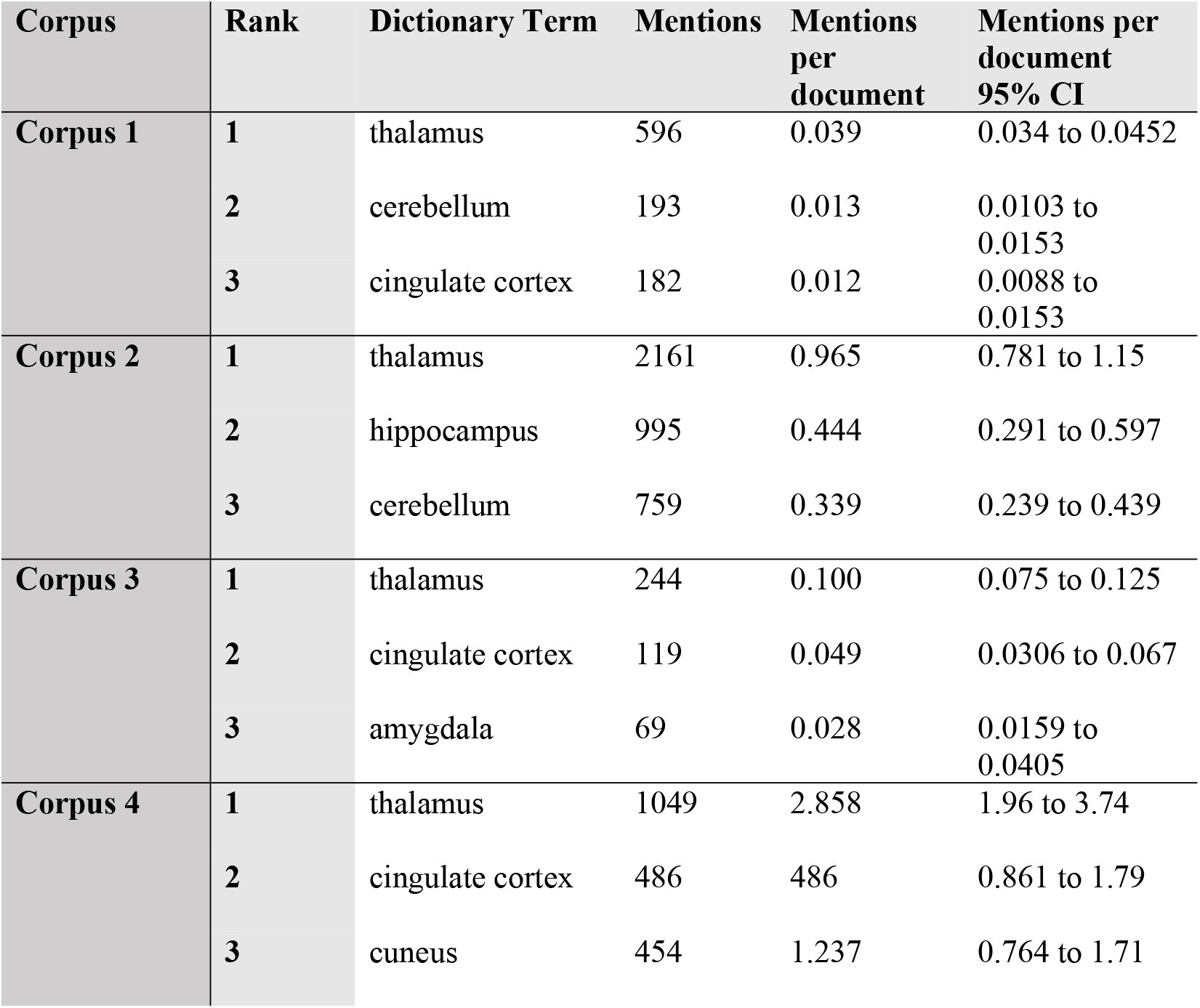
Bibliometric search results highlighting the top 3 highest ranked brain regions in each corpus.

| Corpus | Rank | Dictionary Term | Mentions | Mentions per document | Mentions per document 95% CI |
| --- | --- | --- | --- | --- | --- |
| Corpus 1 | 1 | thalamus | 596 | 0.039 | 0.034 to 0.0452 |
|  | 2 | cerebellum | 193 | 0.013 | 0.0103 to 0.0153 |
|  | 3 | cingulate cortex | 182 | 0.012 | 0.0088 to 0.0153 |
| Corpus 2 | 1 | thalamus | 2161 | 0.965 | 0.781 to 1.15 |
|  | 2 | hippocampus | 995 | 0.444 | 0.291 to 0.597 |
|  | 3 | cerebellum | 759 | 0.339 | 0.239 to 0.439 |
| Corpus 3 | 1 | thalamus | 244 | 0.100 | 0.075 to 0.125 |
|  | 2 | cingulate cortex | 119 | 0.049 | 0.0306 to 0.067 |
|  | 3 | amygdala | 69 | 0.028 | 0.0159 to 0.0405 |
| Corpus 4 | 1 | thalamus | 1049 | 2.858 | 1.96 to 3.74 |
|  | 2 | cingulate cortex | 486 | 486 | 0.861 to 1.79 |
|  | 3 | cuneus | 454 | 1.237 | 0.764 to 1.71 |

**Table 3:** The top 10 highest ranked dictionary terms by Savage score.

| Dictionary Terms | Ranking | Savage Score |
| --- | --- | --- |
| **Thalamus** | 1 | 14.191 |
| **cingulate cortex** | 2 | 8.0243 |
| **hippocampus** | 3 | 6.5231 |
| **Amygdala** | 4 | 6.291 |
| **cerebellum** | 5 | 6.2691 |
| **Cuneus** | 6 | 5.8743 |
| **anterior cingulate cortex** | 7 | 4.8953 |
| **precuneus** | 8 | 4.7636 |
| **insular cortex** | 9 | 3.3331 |
| **posterior cingulate gyrus** | 10 | 2.8231 |

Graphical analysis yielded 4 clusters: cluster 1 had 20 members with the insular cortex [PageRank =0.167] as the most important member; cluster 2 had 29 members with the amygdala [PageRank =0.0199] being most important; cluster 3 had 10 members with the thalamus [PageRank = 0. 0205] being most important; and cluster 4 had 19 members with the cingulate cortex being the most important member [PageRank = 0. 0.020] (Fig 2). An interactive online visualization of this graph can be seen here: https://mango117.github.io/BMedScDOC/network.html along with details of each group included in the supplementary material (Table 4). A 3D visualization of these brain regions in is depicted in Fig 3.

**Table 4.** Distribution **of brain regions between modularity cluster groups in graphical network visualization. Listed in alphabetical order**.

| Graphical Network Modularity Groups |  |  |  |
| --- | --- | --- | --- |
| Group 1 | Group 2 | Group 3 | Group 4 |
| heschls_gyrus | amygdala | lateral_geniculate_thalamus | cerebellar_crus |
| insular_cortex | angular_gyrus | lateral_posterior_thalamus | cingulate_cortex |
| lateral_orbital_gyrus | caudate_nucleus | medial_geniculate_thalamus | cuneus |
| lenticular_nucleus | cerebellum | pulvinar_anterior_thalamus | dorsal_anterior_cingulate_cortex |
| lenticular_nucleus_pallidum | fusiform_gyrus | pulvinar_inferior_thalamus | dorsolateral_prefrontal_cortex |
| lenticular_nucleus_putamen | gyrus_rectus | pulvinar_lateral_thalamus | frontal_eye_fields |
| locus_coeruleus | hippocampus | pulvinar_medial_thalamus | anterior_cingulate_cortex |
| olfactory_cortex | inferior_occipital_gyrus | thalamus | anterior_cingulate_cortex_pregenual |
| perirhinal_cortex | inferior_parietal_gyrus | thalamus_anteroventral_nucleus | lingual_gyrus |
| primary_somatosensory_cortex | inferior_temporal_gyrus |  | anterior_cingulate_cortex_subgenual |
| primary_visual_cortex | middle_cingulate_gyrus |  | orbitofrontal_area |
| raphe_nucleus | middle_frontal_gyrus |  | parahippocampus |
| red_nucleus | middle_occipital_gyrus |  | posterior_cingulate_gyrus |
| retrosplenial_cortex | middle_temporal_gyrus |  | anterior_prefrontal_cortex |
| secondary_visual_cortex | orbital_inferior_frontal_gyrus |  | precuneus |
| substantia_nigra | paracentral_lobule |  | premotor_cortex |
| temporopolar_area | paracingulate |  | primary_motor_cortex |
| ventral_entorhinal_cortex | postcentral_gyrus |  | supplementary_motor_cortex |
| ventral_tegmental_area | precentral_gyrus |  | associative_visual_cortex |
| auditory_cortex | anterior_temporal_lobe |  |  |
|  | rolandic_operculum |  |  |
|  | superior_frontal_gyrus |  |  |
|  | superior_occipital_gyrus |  |  |
|  | superior_parietal_lobule |  |  |
|  | superior_temporal_gyrus |  |  |
|  | supramarginal_gyrus |  |  |
|  | ventral_anterior_cingulate_cortex |  |  |
|  | vermis |  |  |
|  | calcarine_fissure |  |  |

**Fig. 2:**
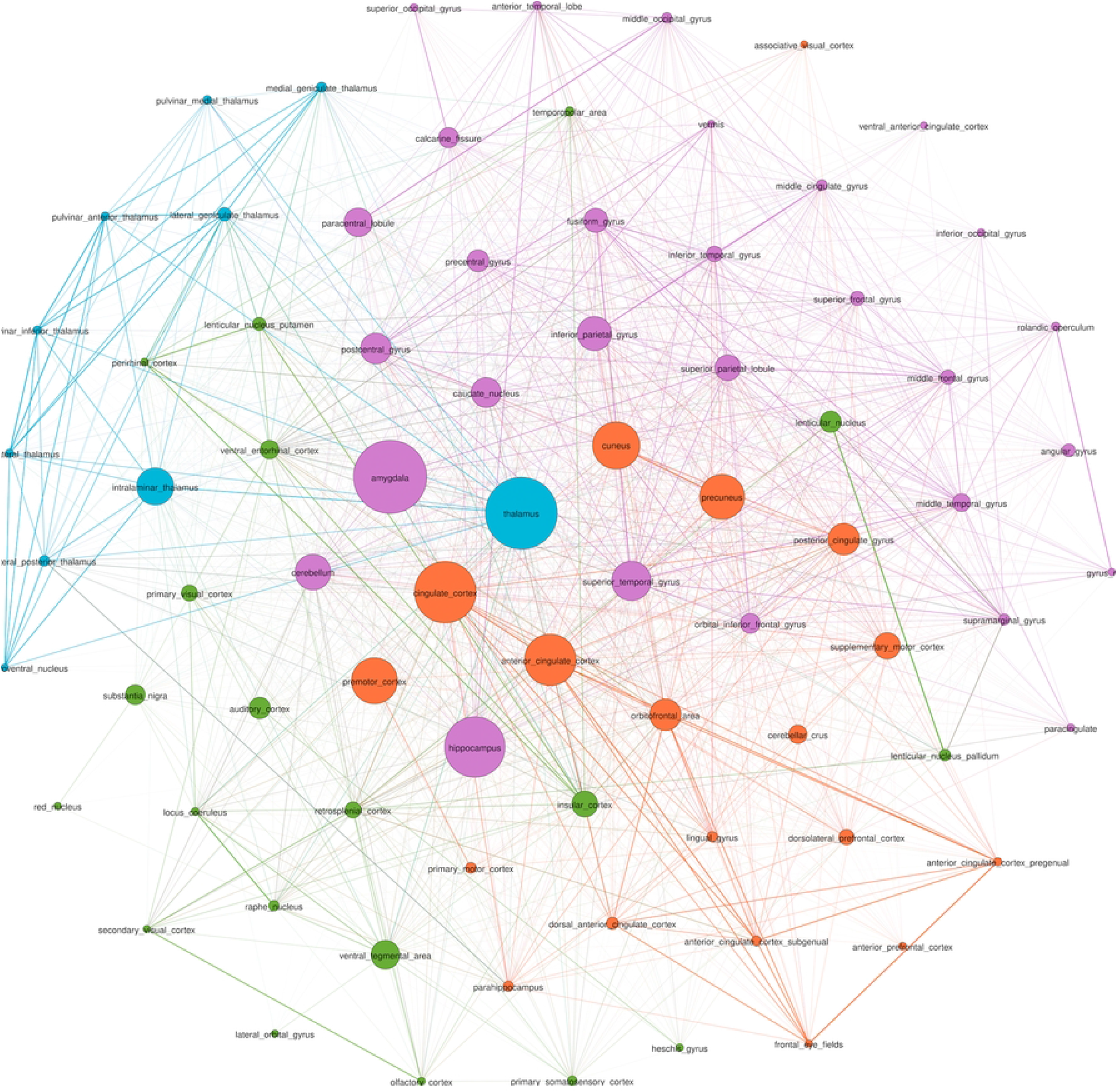
Graph network visualization of dictionary terms found in the full-texts subgroup corpus (Corpus 4). Node sizes are weighted by betweenness centrality, and communities are color coded by modularity cluster. (Green) Cluster Group 1. (Purple) Cluster Group 2. (Blue) Cluster Group 3. (Orange) Cluster Group 4. A description of each group is in Table 4. An interactive online visualization of this graph can be seen here: https://mango117.github.io/BMedScDOC/network.html

**Fig. 3:**
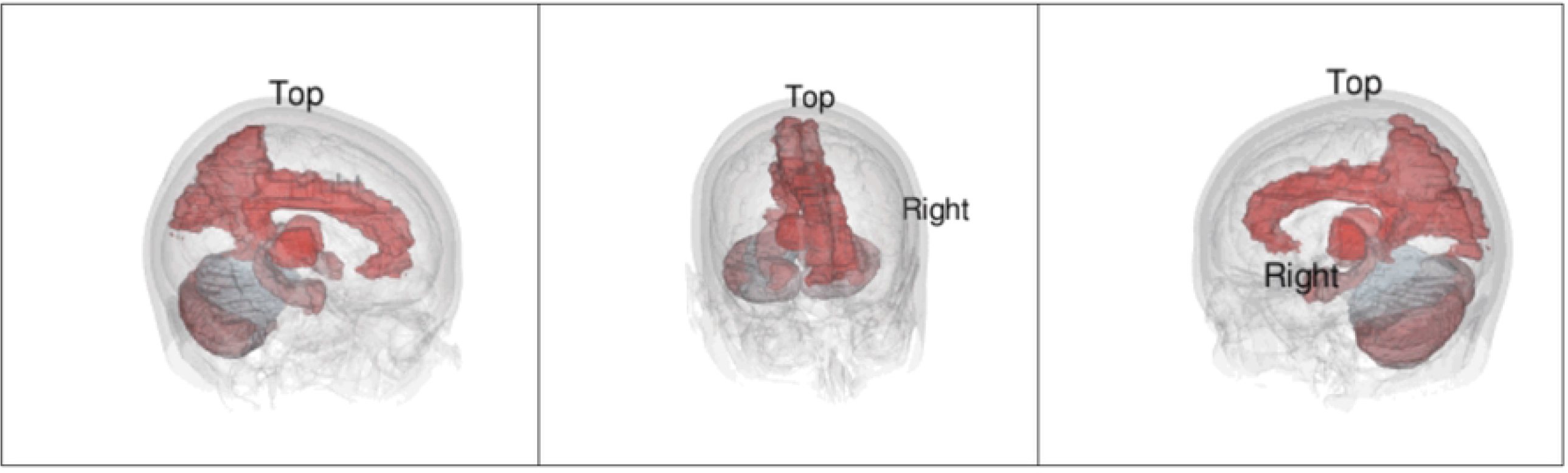
Still images of a 3D visualization of the top 10 most frequently mentioned terms from text-mining of the full-texts corpus. Brain regions are colored in a red to light blue gradient, with the most frequent region being a dark red. An animated 3D GIF of this visualization can be seen here: https://mango117.github.io/BMedScDOC/braingif.html

## Discussion

Our results indicated that the thalamus and cingulate cortex were the most frequently mentioned anatomical brain regions in DoC studies. These findings are not surprising given these brain structures have a well-established link with awareness and arousal in current DoC literature. The novelty of our approach is in taking written text in scientific literature and producing visualization tools for both clinicians and the lay public. In this paper, we were also able to apply graph theoretical analysis to discern the underlying structure of the connections among these brain structures. These findings were performed by collating available techniques and using open-source software and open data; these methods are documented in this paper and the code has been made available on Github. This reproducible approach provides a way for clinicians and other citizen scientists to undertake similar research on this topic or related topics in neuroscience and other fields.

### Anatomical Findings

The high frequency of thalamus mentions per research paper in the full-texts corpora (2.858, 95% CI 1.96 to 3.74) is highly congruent with current literature. The thalamus inhabits the dorsal component of the diencephalon, between the cerebral cortex and midbrain. This location allows it to act as a hub to relay information between subcortical areas and the cerebral cortex.[31] Most sensory inputs are sent to the thalamus before being redirected via many thalamocortical radiations to their final destinations in the outer cortex.[32] This key anatomical feature aligns with our graph theory analysis. The high PageRank score of 0.02046 indicates many highly weighted connections between the thalamus and other nodes in the network, with 96.15% of nodes sharing an edge with it. Its high centrality on the network visualization (Fig 2) is contrasted with the distance between the rest of its modularity cluster (Fig 2, cluster group 3). This indicates that it shares many connections with various other cortical regions rather than being highly connected to a single region of the brain. In the literature, the thalamus is strongly connected with both awareness and arousal in DoC. Awareness relies on high connectivity between the thalamus and cortex, and arousal requires brainstem signals to be forwarded to the thalamus.[33, 34] Its further importance as a treatment target for deep brain stimulation indicates that downregulation of the thalamus likely has considerable implications on conscious state.[35]

The cingulate cortex is another central neurological structure mentioned frequently (PageRank = 0.02022, Savage score = 8.024). It is in the medial aspect of the cerebral hemispheres. It is often categorized into three parts – the anterior cingulate cortex (ACC), the midcingulate, the posterior cingulate cortex (PCC).[36] The ACC (PageRank = 0.0196, Savage score = 4.8953) is often linked to emotion due to dense interconnectedness with limbic system structures such as the amygdala, sharing a strong edge weighting on the graphical visualization (Fig 2). The ACC’s role in self-perception has been previously correlated to the level of consciousness. A fMRI study by Qin et al. illustrated ACC signal changes when DoC patients were exposed to self-related auditory stimuli which further correlated with the patient’s level of consciousness.[37] The PCC (PageRank = 0.0177, Savage score = 2.8231) has previously well-established links with consciousness. The PCC forms a central node in the DMN, active during “wakeful rest.”[16]

More interesting, however, are the findings linking brain regions that are not as commonly associated with DoC. The amygdala (PageRank = 0.01987, Savage score = 6.291) and hippocampus (PageRank = 0.01881, Savage score = 6.5231) are midline areas strongly linked to the limbic system functions of emotional responses and memory.[38] Both share input from the thalamus, as mirrored by the high centrality of all three locations on the graphical visualization (Fig 2).[39] The high frequency of these two regions alongside the ACC may suggest limbic involvement in DoC. This has been highlighted by a few previous studies into pain perception during DoC, but this conclusion cannot be made based on text mining analysis alone.[40-42] The cuneus (PageRank = 0.01956, Savage score = 5.8743) is another midline area. Previous literature has demonstrated an increase in cuneus density associated with increasing conscious state during DoC recovery, but once again, literature on this relationship is sparse.[43-45]

### Comments on Experimental Design

Text mining, sometimes referred to as natural language processing (NLP), is a way of converting unstructured text into structured data.[24] This allows for automated analysis of large corpora that would otherwise be infeasible for a human researcher to analyze. A human reader would have trouble processing, memorizing and understanding 14,945 abstracts and 2178 full-texts on this topic. A domain knowledge expert would have this knowledge having spent many years working in the field. By contrast, a newcomer to the topic would not be able to do this.

In this study, we have illustrated a different approach to text mining scientific literature. In the case of DoC literature, there are different methods between studies due to varying imaging techniques or diagnostic criteria used.{Giacino, 2002 #51}[46-48] Text mining also allows some overlooked details to be picked up on, such as the highlighting of limbic system structures in this study. This is often because of the “objective” analysis a computer is able to perform without any pre-existing ideas or biases regarding a research topic.

### Limitations

There are limitations when using text mining to analyze scientific literature.[49] Many anatomical terms we searched for were long tail keywords (ie. “Left supplementary motor area”) comprised of multiple nouns and adjectives. The linguistic structure of these terms presents a limitation for text mining.[50] In writing, these terms are rarely fully written out by researchers and often have a high degree of variation (i.e. using acronyms or shorthand). It was noted that different publications interchangeably used words such as “region”, “gyrus”, “area” and “cortex” when describing brain regions. We utilized pre-processing of data in each corpus to account for such linguistic idiosyncrasies.[24]

Unlike human researchers, a text mining algorithm is unable to grasp the context of each individual word or document. This is unavoidable in the case of such a large dataset with millions of words, unless a human were to perform this study entirely by hand.

Countermeasures such as subgroup analysis using NMF topic modelling provide an added layer of accuracy to text mining.[21] By separating a large dataset into smaller corpora based on topics, we can compare different subgroups to better understand the corpora holistically. With a large enough dataset, effective pre-processing, and topic modelling, the limitations of text mining can be minimized.

## Conclusion

In this study, text mining of DoC literature revealed the cingulate cortex and thalamus are strongly associated with DoC, likely due to the roles they play in maintaining awareness and comprising the Default Mode Network (DMN). Areas of the brain such as the cuneus, and limbic system regions like the amygdala and hippocampus, were also prominently mentioned in literature. These structures and their relationships are documented in our figures and on the web. Their relationship to DoC should be further investigated.

## Data Availability

The data used for the study is publicly available data from a pubmed eSearch “Disorders of Consciousness” on 9th July 2021. Data and code used in statistical analysis is publicly available in the Github repositories found at the following 2 URLs: https://github.com/Mango117/BMedScDOC_Scripts https://github.com/Mango117/BMedScDOC_Graph

https://github.com/Mango117/BMedScDOC_Scripts

